# Comparison study of commercial COVID-19 RT-PCR kits propose an approach to evaluate their performances

**DOI:** 10.1101/2021.01.20.21250143

**Authors:** Hana Krismawati, Caroline Mahendra, Annabelle Hartanto, Muhammad Fajri Rokhmad, Semuel Sandi, Astrid Irwanto

## Abstract

With the increasing number of COVID-19 cases in Indonesia, scalable and high-throughput diagnostic testing is essential nationwide. Currently, RT-PCR has been the preferred method of viral detection and many manufacturers offer commercial kits for routine clinical diagnostics. In response to the incoming of various kits, there is a need to assess their performance and compatibility of use in clinical laboratories. Kit characteristics impact the testing workflow of these laboratories and some factors can render a kit to perform sub-optimally, leading to false results that are misleading for public safety. Here, we evaluated six commercial kits that are predominantly distributed to appointed testing facilities across Indonesia. Their performance was assessed based on their ease of use, availability, robustness and accuracy for scalable testing in a manual set-up. Our findings demonstrated that all six kits are suitable for use in routine diagnostics, but their considerations for use may vary according to different use-cases. To better guide considerations in procurement of kits, our study provided a systematic approach for laboratories to assess the performance of new incoming kits.

## INTRODUCTION

On 30 January 2020, WHO declared the COVID-19 outbreak a public health emergency of international concern (PHEIC) and by 11 March 2020 it was officially characterized as a pandemic(1). Globally, as of 19 October 2020, there have been 39,944,882 confirmed cases of COVID-19 including 1,111,998 deaths, reported to WHO(2). Indonesia reported their first case of infection on 2 March 2020(3), and the first 11 cases described had partial N gene sequence 100% similarity with the SARS-CoV-2 sequence from Wuhan(4). As of 21 October 2020, the numbers have risen to 373,109 confirmed, 12,857 dead and 297,509 recovered cases(5). WHO has emphasized the importance of timely diagnosis of infected individuals to control the spread, where Nucleic Acid Amplification Tests (NAATs) remain the recommended method for screening current infection(6). Reverse Transcription Polymerase Chain Reaction (RT-PCR) gained popularity worldwide and many manufacturers offer commercial kits for routine clinical diagnostics. In response to the flood of commercial kits in the market, on 28 April 2020 the Indonesian Committee of COVID-19 Task Force released a list of 115 RT-PCR kits recommended for clinical testing(7). The list was created based from WHO guidelines, but is lacking evidence and assessment on their performance. Thus, there is a need to assess their performance and compatibility of use in Indonesian laboratory settings. Aside from their analytical validations, many external factors can render a kit to perform sub-optimally, leading to invalid results that are unfavorable.

A number of comparison studies have been conducted for commercial kits that are readily available. One compared the performance of six commercial kits from various regions, and found that all six can be used for routine diagnostics(8). Other studies have focused to assess kits that are domestically available in respective regions. In Korea, four commercial kits have been released to the market until 15 March 2020 and a study evaluated that all four indicated suitable performance for COVID-19 diagnosis and follow-up testing(9). Another study comparing two domestic kits in Guangxi, China concluded that one was more sensitive than the other(10).

Here, we evaluated six commercial kits that are most commonly distributed to appointed testing facilities across Indonesia. Two of them have been confirmed in the previously mentioned studies as suitable for diagnosis in their domestic regions(9,10). Their performance was assessed based on their rate of viable results, accuracy in virus detection, and capacity for scalable testing in Indonesia. This study aims to provide an evaluation process to guide decisions for procurement of quality and suitable RT-PCR kits to better assist in efficient COVID- 19 diagnostics.

## METHODS

### Clinical Sample Selection

This study was conducted on clinical samples consisting of nasopharyngeal and oropharyngeal swab specimens collected inside a tube containing viral transport medium (VTM) that are transferred to the National Institute of Health Research and Development at Papua. Samples recruited for this study were unlinked to all patient information and reassigned new identification numbers. A panel of clinical samples confirmed positive or negative for SARS-CoV- 2 according to their reported clinical diagnosis were collected to evaluate the clinical performance of kits. A handful of routine clinical samples at a given day sufficient for two runs on a 96-well plate were selected for assessing throughput of the commercial kits. (Figure 1)

**Figure 1.**
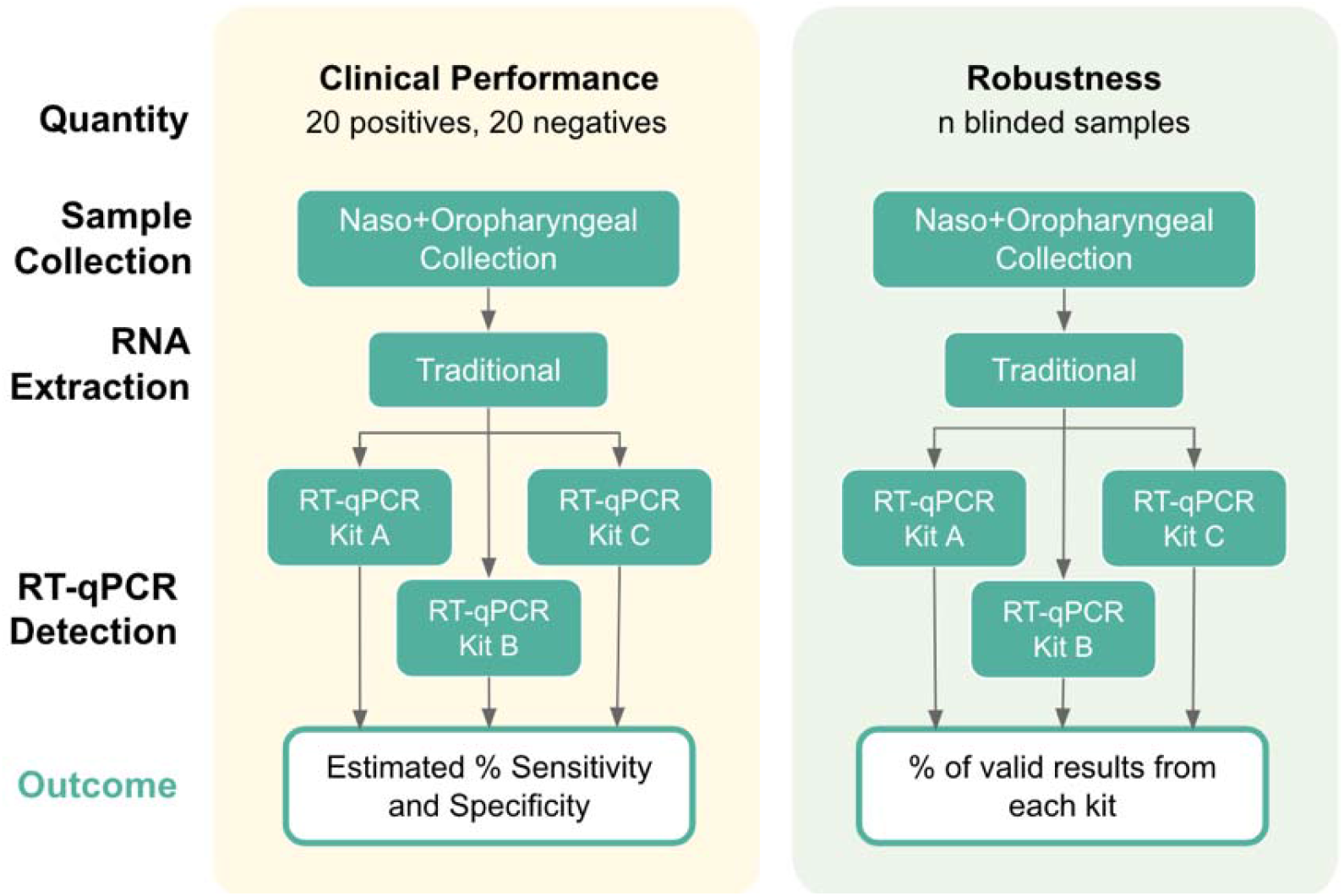
Summary of the workflow for clinical specimens selected for assessing kit performance.

### Selection of Commercial RT-PCR Kits

Commercial RT-PCR Kits were selected based on their availability in the testing facility receiving kits for COVID-19 diagnostics from the Indonesian National Board for Disaster Management, BNPB. All six can be found on the list of kits recommended by the Indonesian Committee of COVID-19 Task Force(7), and their respective specifications derived from their manufacturer’s Instructions For Use (IFU) can be found in Tables 1 and 2.

**Table 1.**
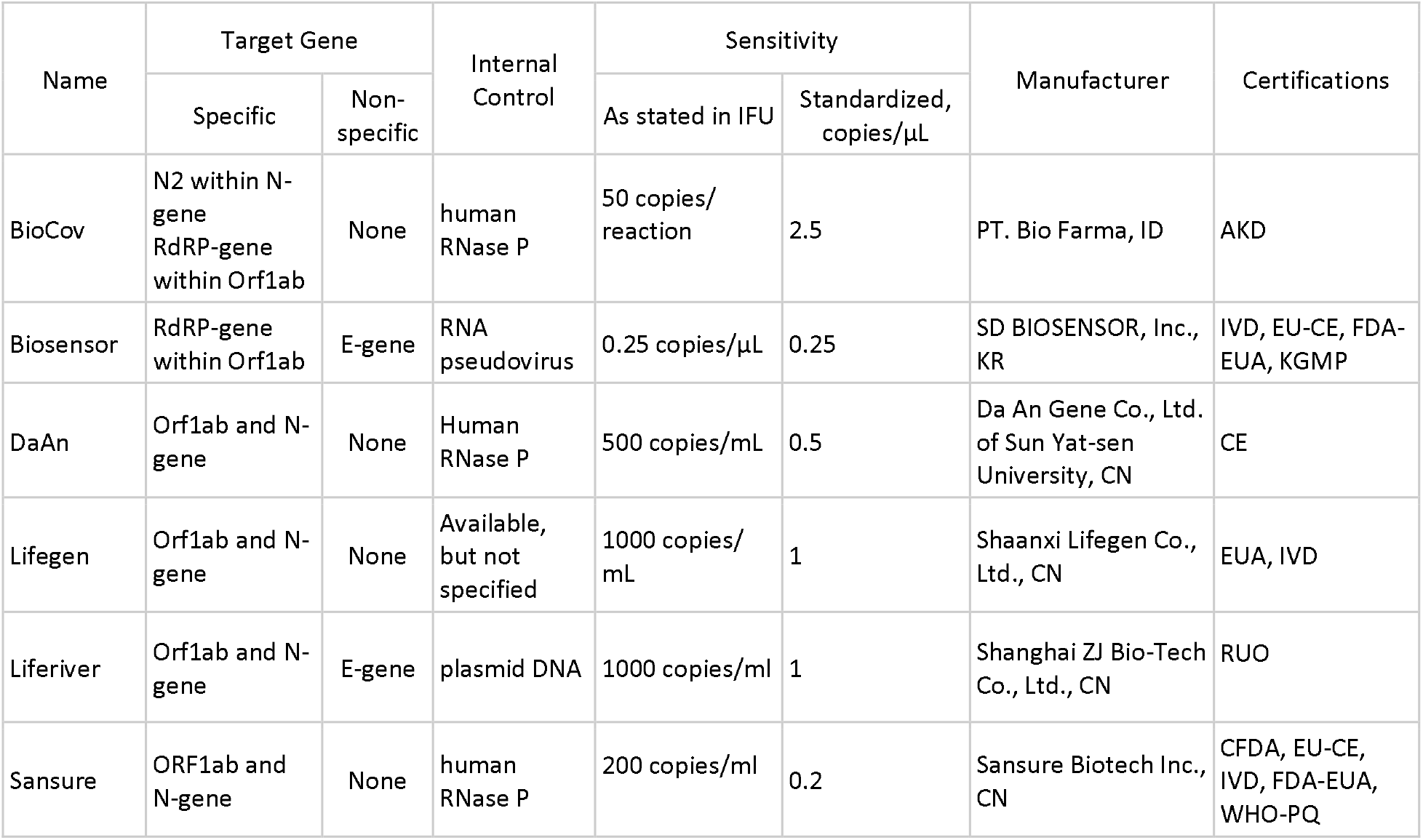
Overview of the six commercial SARS-CoV-2 RT-PCR kits included in this study.

| Name | Target Gene |  | Internal Control | Sensitivity |  | Manufacturer | Certifications |
| --- | --- | --- | --- | --- | --- | --- | --- |
| | Specific | Non-specific | | As stated in IFU | Standardized, copies/μL | | |
| BioCov | N2 within N-gene RdRP-gene within Orf1ab | None | human RNase P | 50 copies/reaction | 2.5 | PT. Bio Farma, ID | AKD |
| Biosensor | RdRP-gene within Orf1ab | E-gene | RNA pseudovirus | 0.25 copies/μL | 0.25 | SD BIOSENSOR, Inc., KR | IVD, EU-CE, FDA-EUA, KGMP |
| DaAn | Orf1ab and N-gene | None | Human RNase P | 500 copies/mL | 0.5 | Da An Gene Co., Ltd. of Sun Yat-sen University, CN | CE |
| Lifegen | Orf1ab and N-gene | None | Available, but not specified | 1000 copies/mL | 1 | Shaanxi Lifegen Co., Ltd., CN | EUA, IVD |
| Liferiver | Orf1ab and N-gene | E-gene | plasmid DNA | 1000 copies/mL | 1 | Shanghai ZJ Bio-Tech Co., Ltd., CN | RUO |
| Sansure | ORF1ab and N-gene | None | human RNase P | 200 copies/mL | 0.2 | Sansure Biotech Inc., CN | CFDA, EU-CE, IVD, FDA-EUA, WHO-PQ |

**Table 2.**
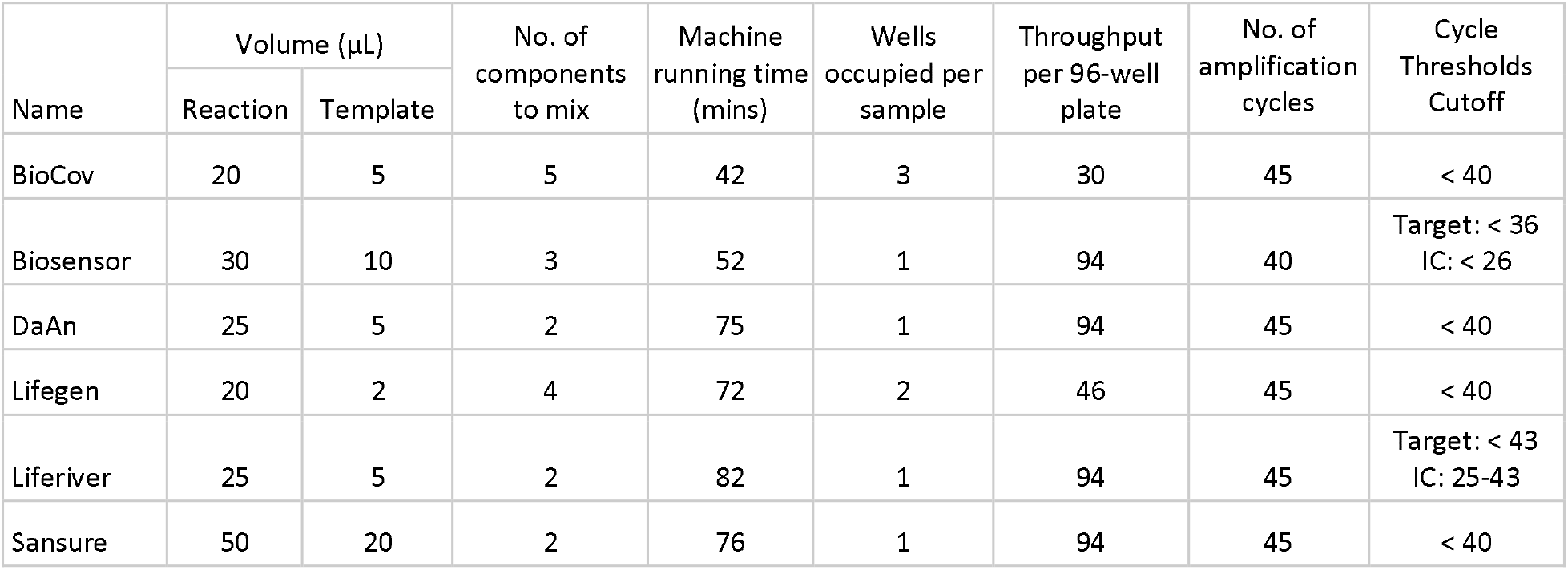
Specifications and characteristics of the six commercial kits, as derived from their respective IFU.

| Name | Volume (μL) | | No. of components to mix | Machine running time (mins) | Wells occupied per sample | Throughput per 96-well plate | No. of amplification cycles | Cycle Thresholds Cutoff |
| --- | --- | --- | --- | --- | --- | --- | --- | --- |
|  | Reaction | Template |  |  |  |  |  |  |
| BioCov | 20 | 5 | 5 | 42 | 3 | 30 | 45 | < 40 |
| Biosensor | 30 | 10 | 3 | 52 | 1 | 94 | 40 | Target: < 36<br>IC: < 26 |
| DaAn | 25 | 5 | 2 | 75 | 1 | 94 | 45 | < 40 |
| Lifegen | 20 | 2 | 4 | 72 | 2 | 46 | 45 | < 40 |
| Liferiver | 25 | 5 | 2 | 82 | 1 | 94 | 45 | Target: < 43<br>IC: 25-43 |
| Sansure | 50 | 20 | 2 | 76 | 1 | 94 | 45 | < 40 |

### Nucleic Acid Extraction

Specimens arriving in the laboratory are stored at -20°C and processed within 24hours. All samples were extracted using AccuPrep^®^ Viral RNA Extraction kit (Ref. K-3033, Bioneer) following the manufacturer’s IFU. Remaining VTM and RNA extract were disposed after the release of test results, unless selected as a panel for evaluation. Samples that were previously diagnosed with a cycle threshold (C_t_ value) less than 25 for SARS-CoV-2 target gene were included for the collection of positives, while those with no detection of target gene as negatives. Selected samples were re-extracted into a total volume of 60 µL and stored as 30 µL aliquots at −80°C.

### Viral Detection through RT-qPCR

RT-qPCR assays were set up following their respective IFU for the six commercial kits as their stated reaction volume and thermocycling conditions. Amplification was performed using Bio-Rad CFX96 Real-Time Thermocycler and read using CFX Manager Version 3.1.

### Data Analysis

Baseline settings and thresholds were applied according to the settings as stated in each kits’ respective IFUs. Results were reported as “Positive”, “Negative”, “Inconclusive” or “Invalid” based on the amplification curve and Ct cutoff as specified in manufacturer’s IFU. “Inconclusive” results were considered when the combination of target genes detected do not confirm positive for SARS-CoV-2 according to the manufacturer’s interpretation guideline. “Invalid” results were reported when there is no evident amplification of target genes or internal control, denoting a failed reaction. During clinical evaluation, results were reported as false when the assessed kit generated results different from the clinical diagnosis for the panel of positive and negative samples. Clinical performance was evaluated by estimating sensitivity and specificity by the following calculation(11):

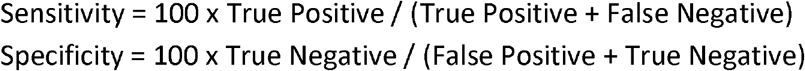

Statistical analyses were conducted using R Version 3.5.2, with p < 0.01 considered as statistically significant.

### Ethical Clearance

This study was approved for ethical clearance by the Health Research Ethics Committee of National Institute of Health Research and Development and expedited with waiver of informed consent. Ethical Approval No.: LB.02.01/2/KE.585/2020

## RESULTS

### Specifications

We first compared the characteristics of the six commercial RT-PCR kits by tabulating specifications derived from their respective Instructions For Use, IFU. Relevant parameters were chosen to assess the ease of use and capacity of high throughput assays (Table 2). Sansure and Biosensor require the largest volume of mix and template added to start a reaction. BioCov and Lifegen recipe contain the most number of components to add into a mastermix, which consequently resulted in the longest time required to set-up. Liferiver, Sansure and DaAn reaction takes the longest to run, since their thermocycling condition requires the highest number of amplification cycles. Finally, while most kits are designed as a multiplex assay for different target genes, BioCov utilizes only one probe color, and Lifegen two. This increased the number of wells occupied per sample to 3-wells for BioCov, one for every target gene and internal control, and 2-wells for Lifegen where each well consist of the specified target gene and corresponding internal control, while the other five kits only occupy 1-well per sample. As a result, both kits have a lower maximum number of samples that can be accommodated per run of a 96-well plate, therefore a lower throughput.

### Clinical Sensitivity and Specificity

To evaluate the clinical performance in accurately detecting SARS-CoV-2, a panel of clinical samples were collected to be tested against the six commercial kits. Out of the total 40 samples, 20 were positive SARS-CoV-2 based on their reported clinical diagnosis and further confirmed when majority of the commercial kits also returned positive for SARS-CoV-2 (Supplementary Table 1). We conducted an ANOVA analysis of Ct value obtained for kits that detect similar target genes for the 20 positive specimens collected for the panel. The independent variables were target gene and commercial PCR kit, where target genes were conditional on the kit being used. We found no statistically-significant difference in average Ct value of target genes detected by different kits (F-value = 0.421, p-value = 0.834). All six commercial kits exhibited statistically similar clinical sensitivity for detection of positive specimens (Figure 2).

**Figure 2.**
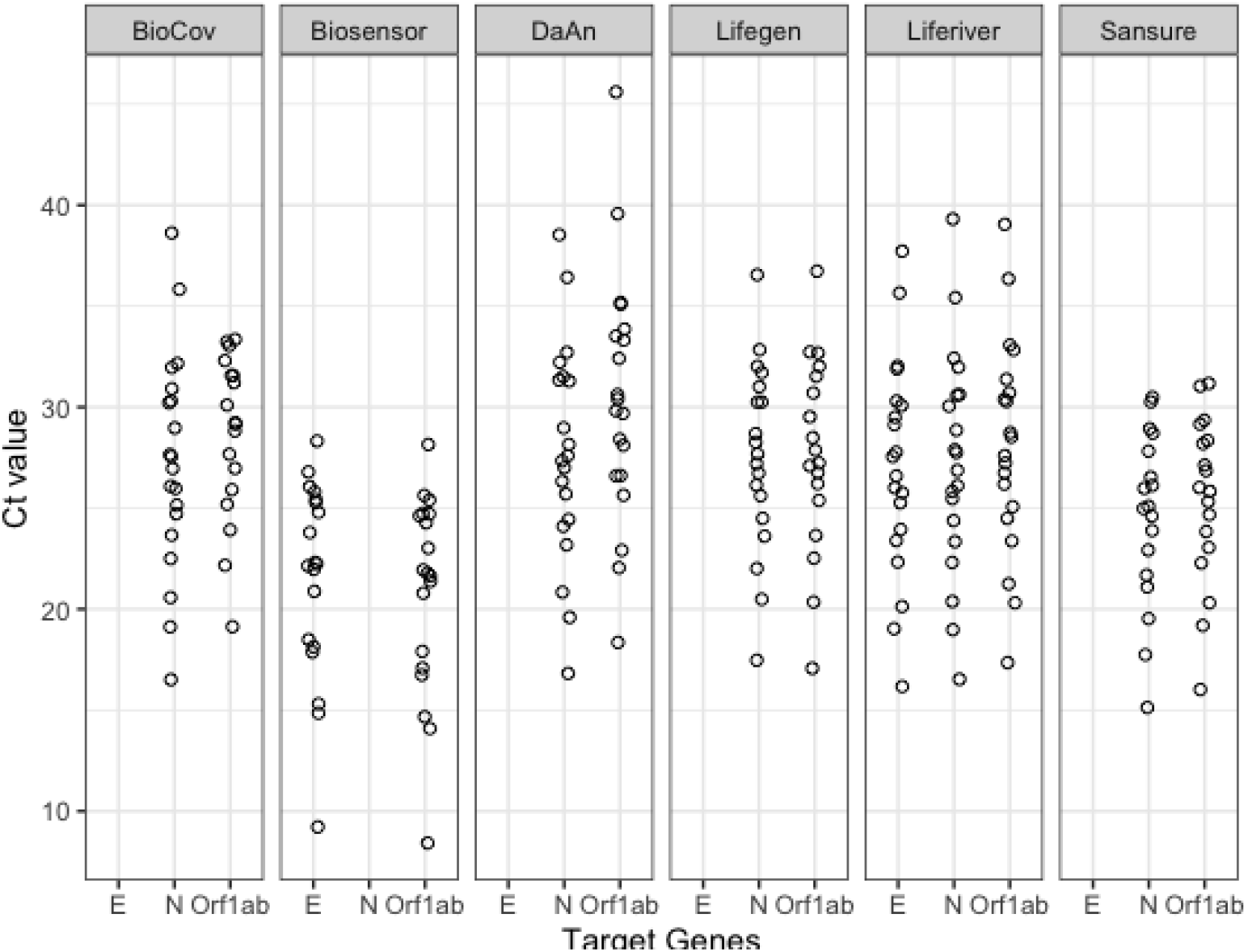
Variation in positive SARS-CoV-2 clinical specimens collected (n=20).

All six kits were able to identify 100% true negatives and 0% false positives, hence demonstrated 100% clinical specificity (Table 3). BioCov, DaAn and Liferiver accurately identified 100% true positives and 0% false negatives, exhibiting 100% clinical sensitivity. Biosensor, Lifegen and Sansure had 95%, 95% and 90% clinical sensitivity respectively, due to detection of false results. Interestingly, one of the false invalid in Biosensor was shared with Lifegen, where it was detected as false negative. Upon closer inspection, this sample was characterized as low positive on BioCov and DaAn, depicted by their late C_t_ value in detection of SARS-CoV-2 target genes. This suggested there may be discrepant result interpretation for this sample, rather than false detection by the different assays. Sansure detected 2 false negatives, which resulted in its lowered sensitivity. Overall, the clinical specificity and sensitivity from the six commercial kits range above 90%.

**Table 3.**
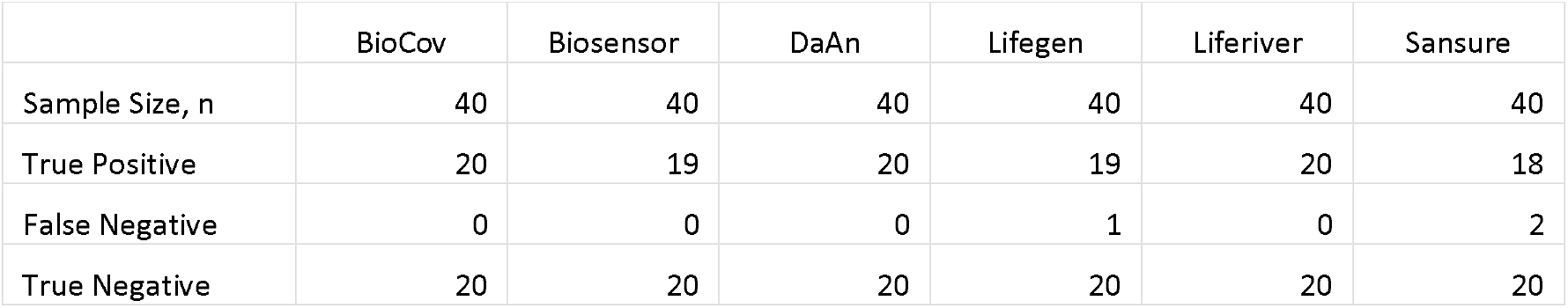
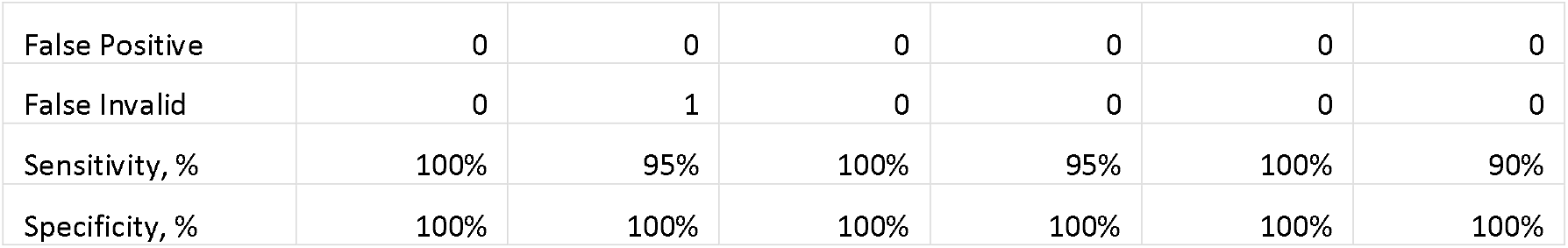
Comparison of results obtained from each kit against their diagnostic accuracy

### Robustness

To assess the robustness of the commercial RT-PCR kits, we subjected each kit to independent runs on clinical samples that were received at a given time (Table 4). Two runs of 96-well plate set-ups were collected for each kit to compare their capacity of throughput testing. All kits were able to optimally utilize the 96-well plate at maximum capacity and obtain sample size of more than 100 from just two runs, except Lifegen and BioCov. Both resulted in sample size less than 70, due to their lower throughput as described in their specifications.

**Table 4.** Comparison of six commercial kits’ robustness

|  | BioCov |  | Biosensor |  | DaAn |  | Lifegen |  | Liferiver |  | Sansure |  |
| --- | --- | --- | --- | --- | --- | --- | --- | --- | --- | --- | --- | --- |
|  | n | % | n | % | n | % | n | % | n | % | n | % |
| Sample size | 60 |  | 157 |  | 148 |  | 67 |  | 101 |  | 105 |  |
| Negative | 48 | 80% | 109 | 69% | 73 | 49% | 55 | 82% | 73 | 72% | 71 | 68% |
| Positive | 12 | 20% | 39 | 25% | 68 | 46% | 10 | 15% | 17 | 17% | 34 | 32% |
| Conclusive | 60 | 100% | 148 | 94% | 141 | 95% | 65 | 97% | 90 | 89% | 105 | 100% |
| Inconclusive<sup>*</sup> | NA | NA | 5 | 3% | NA | NA | NA | NA | 11 | 11% | NA | NA |
| Invalid | 0 | 0% | 4 | 3% | 7 | 5% | 2 | 3% | 0 | 0% | 0 | 0% |
| Repeats | 0 | 0% | 9 | 6% | 7 | 5% | 2 | 3% | 11 | 11% | 0 | 0% |

<sup>\*</sup>Frequency of Inconclusive were obtained only for kits that report "Inconclusive" as stated in their IFU.

Out of the total sample population obtained per kit, all six yielded above 89% “Conclusive” results, which refers to the total number of “Positive” and “Negative” reports obtained when testing using the assessed kits. Samples that were not found “Conclusive” were either “Inconclusive” or “Invalid”, and following laboratory guidelines were subjected for repeat testing. Therefore, “Repeats” refer to the total number of “Inconclusive” and “Invalid” results obtained by the assessed kit. BioCov and Sansure had 0% “Repeats” proving to be the most robust kit at indicating explicit detection of SARS-CoV-2. Lifegen and DaAn generated 3% and 5% “Repeats” respectively, both comprising of all “Invalids”. Biosensor had “Repeats” rate at 6% composed of 3% “Invalid” and 3% “Inconclusive”. Liferiver was the least robust of all with 11% “Repeats” which were all “Inconclusive” results (Table 4).

## DISCUSSION

With the rapid emergence of commercial RT-PCR kits in the market, there is a need for a guided approach to independently assess their performance. Here, we devised a method to evaluate an incoming kit for its suitability in the Indonesian market, where molecular testing is a new field for most medical communities. Local COVID-19 testing facility face constraints primarily from lack of reagents and equipment, and limited human resources(12). It is recommended to start with deriving kit specifications from their respective IFU (Table 1 and 2). This assists in the process of judging a kit’s availability in the market, certifications, throughput, reaction set-up and time required.

Next, it is important to assess their clinical sensitivity, specificity and robustness by conducting benchwork experiments. Clinical sensitivity and specificity were evaluated by subjecting the kit to a panel of clinically diagnosed positive and negative samples. In this study, we collected 20 of each to be retested using the assessed kit and compared their results to their reported clinical diagnosis. However, this number can be minimized to the national standard procedure to validate 10 positives and 5 negatives(13).

Robustness was evaluated by obtaining the rate of valid and conclusive results that a kit generates at any given run. Kits that were tolerant to potential inhibitors are preferred to reduce rate of invalids, while high inconclusive rate were generally seen with kits that also detect the E-gene shared with other lineage of beta-coronavirus(14). In such cases, most kits with this specification advise users to repeat test an inconclusive sample, which can lower throughput when frequent repeat testing is conducted. Highly robust kits minimize the frequency of repeat testing, which can increase the daily testing capacity of a facility.

Following acquisition of the information above, independent kits’ characteristics were listed into a summary table of their assessment results. Table 5 display the parameters considered for evaluating the suitability of a kit for use. Operators favoring high clinical performance and robust kits could consider BioCov with the downside of low throughput and more time spent in setting-up the reaction. Others favoring high capacity of testing and convenient set-up are recommended DaAn which maintained above 95% clinical performance and robustness, but risk longer waiting from machine run-time. Some operators may also choose kits with shorter running time like Biosensor which can reduce the required work hours, while maintaining high throughput and above 94% clinical performance and robustness. Respective laboratories will have their own weighted judgement for each parameter depending on their needs and preferred workflow. Overall, all six kits were competent for use in routine diagnostics of COVID- 19 patients, but their considerations can vary with different use-cases.

**Table 5.** Summary of the commercial kits evaluation

|  | BioCov | Biosensor | DaAn | Lifegen | Liferiver | Sansure |
| --- | --- | --- | --- | --- | --- | --- |
| Certification<sup>a</sup> | IVD | IVD | IVD | IVD | RUO | IVD |
| Reaction set-up<sup>b</sup> | 5 | 3 | 2 | 4 | 2 | 2 |
| Machine time, minutes | 42 | 52 | 75 | 72 | 82 | 76 |
| Throughput<sup>c</sup> | 30 | 94 | 94 | 46 | 94 | 94 |
| Clinical Sensitivity, % | 100% | 95% | 100% | 95% | 100% | 90% |
| Robustness, %<sup>d</sup> | 100% | 94% | 95% | 97% | 89% | 100% |

<sup>a</sup>Regulatory approval obtained from manufacturer's country of origin or at least one country.
<sup>b</sup>As a function of number of components to mix.
<sup>c</sup>As a function of number of samples in a run of 96-well plate
<sup>d</sup>As a function of rate of conclusive results.
Scale and legend of the heatmap displayed below:
Least Preferred Most Preferred

## Data Availability

The datasets presented in this study can be found in online repositories. The names of the repository/repositories and accession number(s) can be found in the article/supplementary material.

## ACKNOWLEDGEMENTS

We thank PT. Nalagenetik Riset Indonesia for assisting in procurement and delivery of the RT-PCR commercial kits to the testing facility. We thank Hutma Hutapea for his supervision at the Institute of Research and Development for Biomedicine Papua in providing and processing clinical specimens included in this study. This study was not endorsed by any kit manufacturer and provides an unbiased and holistic view of all the six commercial kit brands.

## AUTHOR CONTRIBUTION

H.K. and C.M. conceived and designed the study. H.K. and M.F.R. conducted benchwork experiments. C.M. and A.H. performed data analysis and project administration. A.I. and S.S. assisted with study design. C.M. wrote the manuscript with input from all authors.

## CONTRIBUTION TO THE FIELD STATEMENT

In the battle against the COVID-19 pandemic, there has been a flood emergence of commercial RT-PCR kits to support SARS-CoV-2 diagnostics. There is a need for an approach for laboratories to assess kit performance and suitability before adopting any commercial brand. In this study, we compared six RT-PCR kits from different manufacturers on their performance holistically and devised a systematic process to evaluate their suitability for adoption. We aim to provide guidelines to better decision making in procurement of quality RT-PCR kits to assist in efficient COVID-19 diagnostics.

## SUPPORTING INFORMATION

Supplementary Table 1. Evaluation of accuracy of the six commercial kits with previously diagnosed clinical samples

Supplementary Table 2. Results for clinical samples tested using BioCov to assess kit robustness

Supplementary Table 3. Results for clinical samples tested using Biosensor to assess kit robustness

Supplementary Table 4. Results for clinical samples tested using DaAn to assess kit robustness

Supplementary Table 5. Results for clinical samples tested using Lifegen to assess kit robustness

Supplementary Table 6. Results for clinical samples tested using Liferiver to assess kit robustness

Supplementary Table 7. Results for clinical samples tested using Sansure to assess kit robustness

